# Validity of Umbilical Cord Blood Procalcitonin in the Diagnosis of Early-Onset Neonatal Infection

**DOI:** 10.1101/2024.04.30.24306628

**Authors:** Uyen Le Tran, Linh Tran Phuong Giang, Tinh Thu Nguyen

## Abstract

**Introduction:** Early-onset neonatal infection (EONI) poses significant risks to neonatal health, necessitating reliable diagnostic markers for early detection. This study aims to evaluate the diagnostic validity of procalcitonin (PCT) concentration in umbilical cord blood as a biomarker for EONI.

**Methods:** This prospective study was conducted at Ho Chi Minh University Medical Center from April 2022 to September 2022. One hundred eighty neonates with risk factors for EONI were recruited. The PCT level was measured in umbilical cord blood at birth. Based on clinical, laboratory, and microbiologic results, neonates were classified into infected and non-infected groups.

**Results:** Among the neonates studied, 22 (12.2%) were classified as infected and 158 (87.8%) as non-infected. The median PCT in the infected group was significantly higher than that in the non-infected group (0.389 ng/mL vs 0.127 ng/mL, p = 0.007). The optimal PCT cut-off was found by Receiver Operating Characteristic (ROC) to be 0.23 ng/mL, with an area under the curve of 0.87. The results were 59.1%, 98.7%, 86.2%, 94%, 45, and 0.41 for sensitivity, specificity, positive and negative predictive values, positive and negative likelihood ratios, respectively. The post-test probability was 86% if the test was positive and 5% if it was negative.

**Conclusion:** Umbilical cord blood PCT is a reliable marker in the diagnosis of EONI, and its value helps limit the harmful effects of unnecessary prescriptions in non-infected neonates.

## INTRODUCTION

Early onset neonatal infection (EONI) is a major cause of neonatal morbidity and mortality, requiring early diagnosis and treatment to improve outcomes. The diagnosis remains a challenge for pediatricians as the clinical signs are subtle, nonspecific, and develop at a late stage. Moreover, early inflammatory markers such as complete blood cell (CBC) count, high sensitivity C-reactive protein (hs-CRP), and procalcitonin (PCT) are neither sensitive nor specific enough. In contrast, blood cultures, which are the gold standard, are often negative because of low blood volumes drawn and single cultures, as well as the prenatal administration of antibiotics [1]. Because of the reasons mentioned earlier, neonatologists frequently initiate early empirical antibiotic prescriptions before having results of blood cultures and inflammatory markers in neonates who were shown few symptoms or some infection-associated risk factors. Thus, many neonates were subjected to needless treatment. This needless therapy may lead to some serious consequences: risk of the development of resistant bacteria [2], nosocomial infection risk and necrotizing enterocolitis, mother and neonate separation, and increased cost [3–5].

The random-effects estimator for neonatal sepsis incidence in the overall time frame was 2824 cases per 100.000 live births, with a 2.6 times higher incidence of EONI compared to late-onset neonatal infection (LONI), of which an estimated 17.6% died [6]. This estimate was exclusive relying on low - or middle-income country (LMIC) data. Vietnam is one of the LMICs, and the incidence of EONI in full-term neonates in Vietnam was 17 per 1000 live births [7], while this incidence in the United States, one of the high-income countries, was much lower, 0.77 per 1000 live births [8]. Most publications on neonatal sepsis in HICs only include culture-confirmed cases, which means that a positive blood culture establishes a definitive diagnosis of neonatal sepsis. Automated systems for continuous monitoring of blood culture, which are routinely used in the United States, have higher sensitivity and shorten the time to positivity. But in Vietnam, the rate of positive results of blood culture was lower, only 13.8% [7]. Because blood culture testing facilities are not available in most district hospitals and formal healthcare systems prioritize other important healthcare issues, manual blood culture methods have been largely used more than automated systems. Hence, their sensitivity is lower, and the time to positivity is longer.

Due to these challenges and the lack of adequate maternal healthcare infrastructure, such as prenatal screening for GBS, Kaiser early-onset neonatal calculator, NICE, or AAP guidelines are difficult to implement in Vietnam. Therefore, the search for an ideal inflammatory maker in establishing the diagnosis has been ongoing.

In recent years, PCT has been implicated as a sensitive and specific marker of bacterial infection. However, neonatal PCT concentrations undergo a physiologic increase within the first 48 postnatal hours, which complicates the interpretation of results during this period [9]. Therefore, PCT concentrations must be obtained from umbilical cord blood immediately at birth.

This study aims to assess the diagnostic utility of PCT in umbilical cord blood in infants who are at risk of EONI. We hypothesize that the concentration of umbilical cord blood PCT is a reliable marker in EONI.

## METHODS

### Study site

We conducted a prospective study at the Obstetric and Neonatology departments of Ho Chi Minh University Medical Center from April 2022 to September 2022. Ho Chi Minh is the biggest city located in the South of Vienam.

This study was approved by the Institutional Review Board of Ho Chi Minh University Medical Center on June 14th, 2022 (Project No. 2022/46)

### Patients

All neonates delivered during this period were included in the study. Those with risk factors for EONI had a measurement of umbilical cord blood PCT concentration. Additionally, all neonates with risk factors for EONI had drawn blood cultures from cord or venous blood. CBC, CRP, and PCT were determined during the first 24 hours of life and repeated during the first 72 hours of life. They were also examined by the neonatalogist immediately after birth and on subsequent days of life as frequently as required, at least once a day, to detect clinical signs of infection. These neonatalogists, who were blinded for the results of umbilical cord blood PCT concentration, classified neonates into two groups: the infected group and the non-infected group. Therefore, PCT levels were not involved in the diagnosis and clinical decisions.

#### Inclusion criteria

were neonates presenting at least one of the following risk factors for EONI. These risk factors include maternal intrapartum fever of more than 38°C, preterm delivery before 35 weeks gestation, prolonged rupture of membranes (>18 hours), or confirmed maternal colonization by *Group B Streptococcus* with incomplete prophylaxis (<4 hours), or amniotic fluid of abnormal color (green, yellow or meconium-stained) or being malodorous.

#### Exclusion criteria

were death, neonates being transported to another hospital within the first 3 days of life, severe malformation, the missing value of umbilical cord blood PCT, or parent consent not being granted.

Two groups were identified as infected and non-infected based on the clinical, biochemical, and microbiological findings, as well as in compliance with international guidelines [10]. A proven infection was defined by a positive blood culture in the presence of clinical signs or symptoms of infection. A probable infection was defined by the presence of clinical signs and symptoms of infection and at least two abnormal laboratories accompanied by a negative blood culture. Neonates with proven or probable infection were classified as infected. Neonates were considered non-infected in the absence of clinical signs and symptoms of infection and abnormal laboratory results. Categories for neonatal infection are shown in Table 1.

### Data collection

Perinatal data included maternal temperature, time of membrane rupture, the results of the mother’s GBS screening, characteristics of amniotic fluid, and antenatal antibiotic exposure. Clinical information included birth weight, gestational age at birth, gender, delivery method, clinical signs of infection during the first 72 hours of life, blood culture, white blood cell, neutrophil, platelet count, CRP, and umbilical cord blood PCT.

A dosage of umbilical cord blood PCT, obtained by umbilical artery puncture directly after the birth of the placenta in standard vacuum blood collection tubes containing lithium heparin, was systematically realized when the technique was available. The tubes were carried to the biochemistry laboratory within 1 hour. We used the Elecsys BRAHMS PCT assay for quantitative determination of PCT. This assay requires a minimum sample volume of 30 μl and the reaction time is about 18 minutes. The measuring range of the assay was 0.02-100 μg/L with an automated dilution extending the upper range to 1000 μg/L.

### Statistical analysis

All data were analyzed using Stata software version 12.0. We described qualitative data as percentages, accessing precision by means of 95% confidence intervals (CIs) and quantitative data using percentages, medians, and standard deviations. Results were considered statistically significant at P values < 0.05.

We analyzed PCT distribution among infected and non-infected groups. These distributions were normal, so we used the *t*-test to compare umbilical cord blood PCT concentrations between the two groups. The sensitivity, specificity, negative and positive predictive values, negative and positive likelihood ratios of PCT concentration in cord blood were assessed using the receiver operating characteristic (ROC) curve and the area under the curve (AUC). We used Youden’s Index to detect the best cut-off value that resulted in the highest sum of sensitivity and specificity. Post-test probabilities were determined using the Fagan nomogram.

## RESULTS

During the period under study, a total of 1,455 neonates were delivered at our hospital; 200 possessed risk factors for EONI. Twenty neonates were not included; four of these were transported to another hospital, and 16 were missed the value of the cord blood PCT. The final sample consisted of 180 neonates with risk factors for infection. Twenty-two neonates had a proven or probable infection. 158 neonates were considered non-infected. The incidence of infection in neonates presenting with risk factors for EONI was 12% (22/180). The incidence of infection in the whole population was 15 per 1000 live births, as shown in the flow chart of the study in Table 1. Blood culture was positive in three cases (13.6%). The infectious agents were Group B Streptococcus in two cases (66.7%) and Staphylococcus aureus in one case (33.3%). The baseline characteristics of the population are described in Table 2.

### Cord blood PCT value

The median PCT of the infected group was significantly higher than that of the non-infected group (0.389 ng/ml vs. 0.087 ng/ml, p< 0.05) (Table 3). We analyzed the ROC curve and identified a cut-off of 0.23 ng/ml as the optimal compromise, accompanied by an area under the curve of 0.87, a sensitivity of 59.1%, and a specificity of 98.7% (Figure 2). The positive and negative predictive values were 86.2% and 94.7%, and the positive and negative likelihood ratios were 45 and 0.41. Cord blood PCT levels were below 0.23 ng/ml in 165 neonates (91.6%); two of these (1.1%) developed sepsis. PCT levels were at or above 0.23 ng/ml in 15 neonates (8.4%); 13 of these (7.2%) had infection, proven in two of these (1.1%) and probable in 11 cases (6.1%).

**FIGURE 1:**
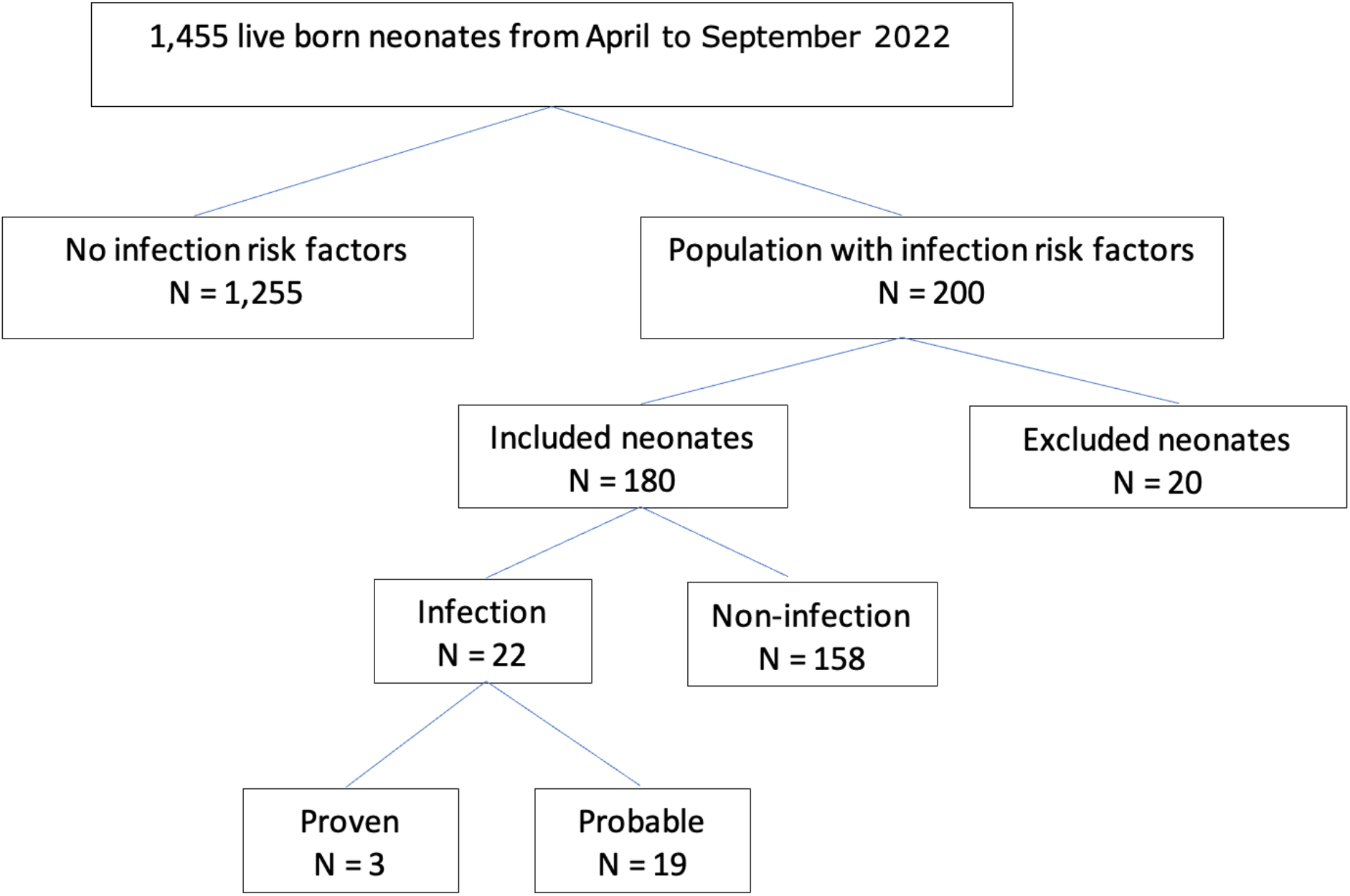
Flow chart displaying the recruitment process and excluded patients in the study.

**FIGURE 2:**
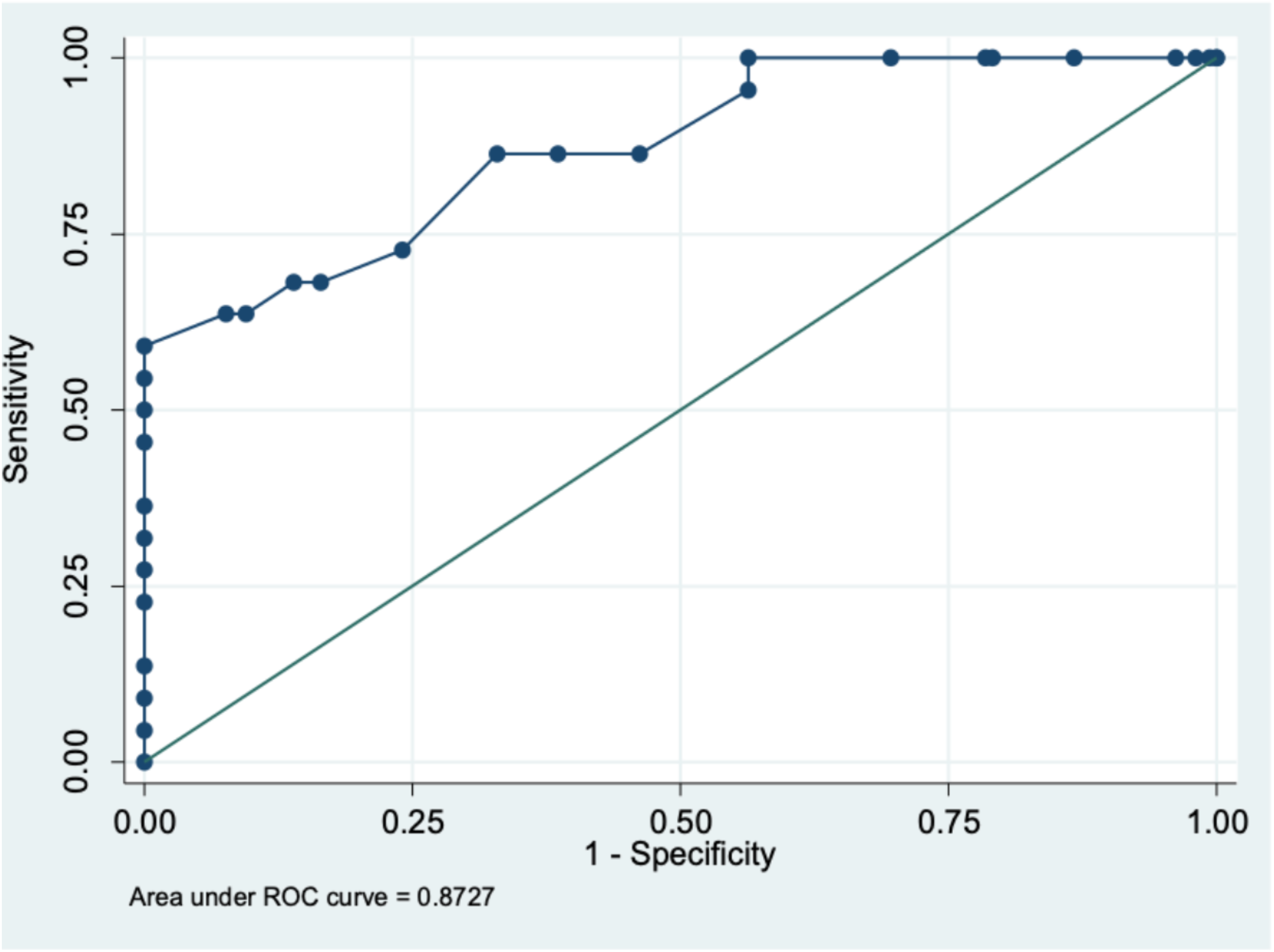
Receiver operating characteristics (ROC) curve of umbilical cord blood procalcitonin.

Figure 3 shows corresponding post-test probabilities determined with the Fagan nomogram. With a 12% prevalence (pretest probability) of EONS, a LR+ of 45 translates into a positive predictive value (post-test probability) of 86%. Similarly, a LR-of 0.41 translates into a 5% post-test probability. These results indicate that, with at least one risk factor present, the probability of infection was 86% when the test was positive and 5% when the test was negative.

**FIGURE 3:**
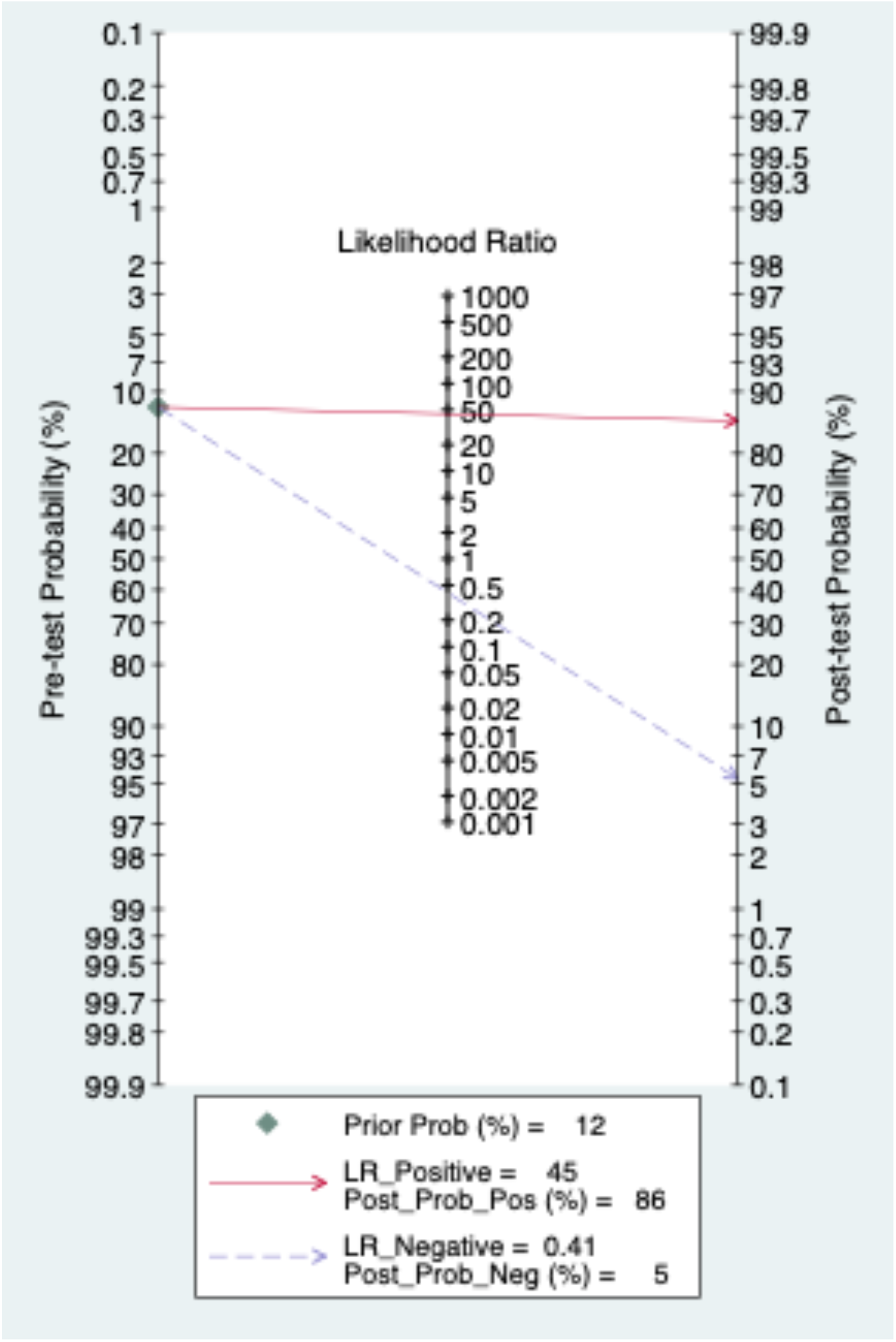
Fagan nomogram representing post-test probability based on positive and negative likelihood ratios.

## DISCUSSION

Neonatal infection, especially early-onset neonatal infection, is a life-threatening condition. Therefore, reducing the burden of EONI is an international medical priority. However, non-specific signs and symptoms make the diagnosis of EONI difficult. New laboratory techniques are needed in early diagnosis.

In recent years, PCT has been reported as a sensitive parameter for early diagnosis. After injecting endotoxin, the procalcitonin concentration rose and was detectable at 4 hours, peaked at 6 hours, and maintained a plateau through 8 and 24 hours [11]. This kinetics in neonate is assumed to be similar to the results of the above study, we expect that the concentration of PCT in cord blood would be abnormal in the case of intrauterine infection. In our study, the median cord blood PCT of the infected group was significantly higher than that of the non-infected group. The PCT concentrations were found to be variable, although most of the values range between 0 and 0.6 ng/ml, as also previously reported. The selected threshold of cord blood PCT varies between studies, depending on factors such as the population study, incidence of EONI, and the evaluating criteria. In our study, the optimal PCT cut-off value was 0.23 ng/ml with sensitivity and specificity of 59.1% and 98.7%, respectively. With a high positive predictive value of 86.2%, PCT is an early and reliable marker in the diagnosis of EONI.

Another noteworthy finding is the capacity of this marker to identify neonates who were non-infected among those with risk factors for infection. According to the Fagan nomogram, a neonate with a risk factor and PCT value below 0.23 ng/ml had a statistical risk of being infected of 5%. Its good negative predictive value (94.7%) results in a reduction in the number of neonates being treated unnecessarily. Therefore, non-infected causes of respiratory distress or hypotension should be aggressively sought if cord blood PCT is negative, especially in preterms. This reduction in antibiotic treatment would represent a direct advantage for neonates because of the potential toxicity of antibiotics and an indirect ecological advantage by reducing antibiotic selection pressure for physicians. Hospitalization for antibiotic treatment of neonates results in the separation of mother and child during these important first days of life and increased cost. Neonates need punctures for new intravenous lines frequently, so reducing antibiotic treatment will result in fewer punctures. To address the problem of increasing global antibiotic resistance rates, the WHO has emphasized the urgent need for enhanced antibiotic stewardship [12]. Each dose of antibiotic treatment takes into account the emergence of antibiotic resistance and changes the human microbiome [13,14]. These changes in the microbiome in early life contribute an important role in shaping the immune system and future health of the individual [15]. Therefore, reducing unnecessary antibiotic treatment to lower the emergence of antibiotic resistance.

The limitation of our study is its low sensitivity. Among 22 infected neonates, only 13 neonates (59%) had a PCT value higher than 0.23 ng/ml, which is lower than that of other studies [16–19]. Recently, a meta-analysis including 17 reviews and 2197 episodes of suspected neonatal infection reported the sensitivity and specificity of umbilical cord blood PCT in diagnosis being 82% and 86% [20]. The first reason was the low incidence of EONI (22/1455), and the small number of cases in the infected group. This small number of infectious cases was responsible for the lack of power and subsequent lack of precision in diagnostic value measurement with a large 95% CI. Moreover, 16/200 neonates (8%) presenting with risk factors for EONI were excluded from this study because cord blood PCT was not accessed. The second reason was that some probable infected cases in the infected group were not really infected. Indeed, the rate of preterms in the infected group was 50% (11 cases). Most of these had respiratory distress syndrome, increased CRP, and antibiotic treatment, so they met the criteria to be finally classified as cases of probable infection. Respiratory symptoms of EONI are non-specific and may overlap with non-infectious causes like RDS in preterms and transient tachypnea. Therefore, these cases were not actually infected.

## CONCLUSION

Our finding suggests that the umbilical cord blood PCT measurement might be a reliable marker for identifying non-infected neonates from those who exhibit risk factors for infection. The approach as mentioned above, has the potential to significantly reduce unnecessary antibiotic prescriptions in the first days of life. Further research is necessary to integrate this biomarker into clinical practice fully.

## Ethics consideration

This study received ethical approval from the Institutional Review Board (IRB) of Ho Chi Minh University Medical Center on June 14th, 2022 (Project No.: 2022/46).

## Informed Consent

All informed consents were obtained, and the data were de-identified. Patient data was anonymized and de-identified throughout the analysis.

## Data Availability Statement

The data presented in this study are available on request from the corresponding author. The data contained within this article is not available due to privacy issues.

## Author contributions

Conceptualization: ULT, LTPG, TTN.

Data acquisition: ULT,.

Analysis and interpretation: ULT, LTPG, TTN.

Supervision: TTN.

Writing draft manuscript: ULT, TTN.

Review, edit, and approve the final manuscript: all authors.

